# The Epilepsy-Cog study: methods to establish a harmonized study of late-onset epilepsy in a meta-cohort of six population-based cohorts in the United States

**DOI:** 10.64898/2026.01.30.26345233

**Authors:** Hyunmi Choi, Jose Gutierrez, Tian Wang, Minghua Liu, Cheng-Shiun Leu, Sylwia Misiewicz, Jiying Han, Natalie Bello, Mary L. Biggs, Emily M. Briceño, Adam M. Brickman, James F. Burke, Ligong Chen, Lisandro D. Colantonio, Stefany P. Diaz Andino, Mitchell S.V. Elkind, Annette L. Fitzpatrick, Christopher Gonzalez Corona, Alden L. Gross, Lei Huang, Emily L. Johnson, W. Craig Johnson, Deborah A. Levine, W.T. Longstreth, Sofia Pelagalli Maia, Richard P. Mayeux, Brian C. Petersen, Oluwadamilola Obalana, Dolly Reyes-Dumeyer, Tatjana Rundek, Danurys Sanchez, Steven J. Shea, Kevin Strobino, Carolyn W. Zhu, Evan L. Thacker

## Abstract

**Objectives:** With the expected demographic shift toward those ≥65 years of age in the United States, late-onset epilepsy (LOE) poses a significant public health issue, yet it has been historically understudied. We are undertaking an effort in the Epilepsy-Cog study to pool individual participant data from six US-based prospective cohort studies. In this paper, we outline the process for ascertaining epilepsy, harmonizing, and pooling individual participant data across the six cohorts.

**Methods:** The Epilepsy-Cog study includes individual participant data from six US-based longitudinal cohort studies: ARIC, CHS, MESA, NOMAS, REGARDS, and WHICAP. In all cohorts except NOMAS, prevalent and incident epilepsy were ascertained using Medicare claims-based algorithms. In NOMAS, epilepsy cases were identified through cohort-based reporting and medical record review. To perform cross-cohort harmonization of variables, we used the lowest common denominator approach, assigning response categories or value levels in common across all cohorts.

**Results:** From a total of 68,544 participants across six cohorts, 43,753 participants met eligibility criteria for Epilepsy-Cog. Among them, we identified 551 (1.3%) participants with prevalent epilepsy and 1,500 (3.4%) participants with incident epilepsy. We have harmonized demographic characteristics, health behaviors, vascular risk factors (VRFs), one genetic variable, medication use, subjective health status measures, incident events, and cause-of-death variables.

**Conclusion:** The Epilepsy-Cog pooled cohort of 43,753 participants with and without epilepsy, combined with harmonized demographic, VRFs, and event data, offers a unique resource to yield new insights into LOE.

## INTRODUCTION

Epilepsy is a common neurological condition affecting older individuals.^1^ The incidence rate increases progressively with age, with the steepest rise occurring in adults aged 65 and older.^2^ Given the expected demographic shift in the United States, with those 65 years or older increasing from 58 million in 2022 to 82 million in 2050,^3^ late-onset epilepsy (LOE) poses a significant public health issue. However, despite the rising burden of disease, LOE has been a historically understudied topic.

Within the past decade, several groups have leveraged well-known population-based cardiovascular cohort studies, such as the Framingham Heart Study (FHS), the Atherosclerosis Risk in Communities Study (ARIC), and the Cardiovascular Health Study (CHS), to perform secondary data analyses of the risk factors associated with LOE, individually within each cohort.^4–6^ These studies have shown that certain vascular risk factors (VRFs), such as hypertension and diabetes, are significant contributors to the development of LOE. Additionally, in CHS, the effects of VRFs on cognitive decline were more potent in the presence of epilepsy than in those with VRFs alone or in those with epilepsy alone.^7^ However, despite the large cohort sizes and population-based design, findings have not been consistent across these studies. The lack of concordance of findings could be due to insufficient statistical power, as the number of LOE cases was considerably smaller in FHS and CHS than in ARIC.

One way to resolve conflicting results across studies is to perform a pooled analysis after harmonizing individual participant data.^8^ A pooled analysis using individual participant data could (a) enhance statistical power, precision, and reduce bias,^8^ and (b) allow advanced modeling, interactions, non-linear associations, and subgroup analysis,^9, 10^ potentially leading to new insights into LOE pathophysiology. A multi-cohort pooled dataset of epilepsy would address the gaps identified in the 2012 Institute of Medicine Report on Epilepsy, which recommended the creation of large-scale longitudinal studies of epilepsy.^11^

Because no single cohort study has been large enough to adequately examine LOE and its consequences, we are undertaking an unprecedented effort in the study Epilepsy-Cog, supported by the National Institute on Aging, to pool individual participant data from six US-based prospective cohort studies. Our goal is to test the hypothesis that LOE represents a clinical manifestation—a marker—of an excess burden of pre-epilepsy VRFs. In future analyses, we will examine the association between LOE, pre-epilepsy VRFs, and cognitive decline. Harmonization of longitudinal cognitive assessments and MRI measures will be detailed in future manuscripts.

Building on harmonization methods previously applied by our co-investigators in studies of blood pressure and cognition across multiple cohorts,^12^ we outline the process we used to pool individual participant data across the six cohorts, including (1) the epilepsy case ascertainment process, grounded in our prior single-cohort work, and (2) the iterative process of harmonizing and pooling of VRFs and incident events, such as stroke, myocardial infarction, and death.

## METHODS

### Study design, setting, and participants

The Epilepsy-Cog pooled cohort study includes individual participant data from six US-based longitudinal cohort studies (Table 1): Atherosclerosis Risk in Communities (ARIC) Stidy,^13^ Cardiovascular Health Study (CHS),^14^ Multi-Ethnic Study of Atherosclerosis (MESA),^15^ Northern Manhattan Study (NOMAS),^16^ Reasons for Geographic and Racial Differences in Stroke (REGARDS),^17^ and Washington Heights/Inwood Columbia Aging Project (WHICAP).^18^ The Columbia University Institutional Review Board (IRB), which serves as the single IRB for Epilepsy-Cog, approved the use of de-identified data from the six cohorts for harmonization and pooling. IRBs of participating institutions involved in the six cohort studies approved the individual cohort studies. All cohort study participants provided written informed consent at the time of cohort entry. The six cohorts were originally established by sampling participants from various geographic locations throughout the United States between the 1980s and the 2000s to study midlife and late-life risk factors for cardiovascular disease, stroke, and cognitive outcomes. Participants in each cohort study have undergone periodic in-person and phone follow-up to ascertain cardiovascular and brain health events such as stroke and dementia, and to update vascular risk factor information.

**Table 1.** Characteristics of six US population-based longitudinal cohorts for Epilepsy-Cog pooled cohort study of epilepsy in late life.

| Characteristic | ARIC | CHS | MESA | NOMAS | REGARDS | WHICAP |
| --- | --- | --- | --- | --- | --- | --- |
| *Original cohorts* |  |  |  |  |  |  |
| Original purpose | Midlife risk factors for atherosclerosis, coronary heart disease, and stroke | Late life risk factors for coronary heart disease and stroke | Midlife and late life subclinical cardiovascular disease prevalence, progression, and risk factors | Midlife and late life risk factors for incident stroke and cognitive decline | Geographic and racial differences in midlife and late life incident stroke and cognitive decline | Aging and late life risk factors for incident dementia and cognitive decline |
| Locations | Washington Co, MD; Minneapolis, MN suburbs; Jackson, MS; Forsyth Co, NC | Sacramento Co, CA; Washington Co, MD; Forsyth Co, NC; Pittsburgh, PA | Los Angeles Co, CA; Chicago, IL; Baltimore Co, MD; St. Paul, MN; New York, NY; Forsyth Co, NC | Northern Manhattan, New York, NY | 48 US States (all except AK and HI), including urban and rural areas | Northern Manhattan, New York, NY |
| Cohort entry years | 1987-1989 | 1989-1990; 1992-1993 | 2000-2002 | 1993-2001; 2001-2008 | 2003-2007 | 1992; 1999; 2009 |
| Sampling frame or method | Probability sampling from state identification cards, or lists of eligible for jury duty | Medicare eligibility lists | Random digit dialing; referrals from participants | Random digit dialing; some household members of participants | Commercially available lists | Medicare eligibility lists |
| Exclusion criteria | Planning to move within 6 months; younger than age 45 and older than age 64 | Being institutionalized or home-bound; receiving cancer treatment; planning to move within 3 years; unable to give informed consent | Known clinical cardiovascular disease; younger than 45 or older than 84; living in a nursing home or on a waiting list; plan to move within 5 years; race/ethnicity other than African American, White, Chinese, or Hispanic | History of stroke; resided in northern Manhattan for less than 3 months; younger than 40 years of age | Race other than African-American and white; active cancer treatment; medical conditions preventing long-term participation; being institutionalized; inability to communicate in English; younger than 45 years | Severe cognitive problems or diagnosed dementia; inability to communicate in English or Spanish |
| In-person follow-up | Every three years through 1998; irregular intervals 2011 to present | Annually 1990-1999; once in 2005-2006 | Every 18-24 months 2000-2007; once 2010-2012; once | When screened positive for incident stroke by phone | Once in 2013-2016 | Every 18-24 months through the present |
|  |  |  | 2016-2018; and<br>once 2022-2024 |  |  |  |
| Phone follow-up | Annually prior to<br>2011 and semi-<br>annually to present | 6-month intervals<br>through 2023 | Annually to the<br>present | Annually until 2024 | 6-month intervals to<br>the present | If participants are<br>unable to return for<br>in-person visits,<br>phone visits are<br>attempted |
| Age range at<br>cohort entry | 45-64 | ≥65 | 45-84 | ≥40 | ≥45 | ≥65 |
| Cohort sample<br>size | 15,792 | 5,534* | 6,814 | 3,497 | 30,239 | 6,668* |
| *Medicare claims*<br>*linkage for epilepsy*<br>*case ascertainment* |  |  |  |  |  |  |
| Years of claims<br>availability | 1991-2018 | 1992-2019; earlier<br>than 1992 for some<br>participants | 2000-2019; earlier<br>than 2000 for some<br>participants | N/A | 2000-2020 | 1999-2019 |
| Claims-linked<br>sample size | 14,702 | 5,256 | 5,522 | N/A | 20,403 | 4,600 |

Abbreviations: ARIC, Atherosclerosis Risk in Communities; CHS, Cardiovascular Health Study; Co, county; FFS, fee-for-service; NY-SPARCS, New York Statewide Planning and Research Cooperative System; MESA, Multiethnic Study of Atherosclerosis; NOMAS, Northern Manhattan Study; REGARDS, Reasons for Geographic and Racial Differences in Stroke; WHICAP, Washington Heights/Inwood Columbia Aging Project.
\* While CHS enrolled 5,888 participants, we considered only the 5,534 participants who were alive at the 1992/93 exam because CHS Medicare linkage began in 1992 for most participants. While WHICAP enrolled 6,792 participants, we considered only the 6,668 participants who were not also enrolled in NOMAS.

ARIC, CHS, MESA, REGARDS, and WHICAP were linked to Medicare claims (Figure 1),^19–22^ which we used to ascertain prevalent and incident epilepsy cases (see below). Linkage to Medicare claims was necessary to determine epilepsy status since these cohorts were not designed to capture epilepsy as an exposure or outcome. Medicare insures more than 98% of U.S. adults aged ≥65, providing a unique opportunity to use claims-based algorithms to identify epilepsy in this population. For the five Medicare-linked cohorts, eligibility to enter Epilepsy-Cog required participants to (a) have enrolled into the cohort study, (b) be linked to Medicare claims, (c) fulfill a minimum of two years of continuous enrollment in Medicare fee-for-service (enrolled in both Parts A and B without being enrolled in Part C [A+B−C], with no gap in A+B−C enrollment longer than one month), and (d) be 65 years of age or above at the two-year mark of continuous Medicare enrollment or the date of cohort enrollment, whichever occurred later. The requirement that participants be 65 years of age or older ensured that their eligibility for Medicare was based on age rather than on disability.

**Figure 1.**
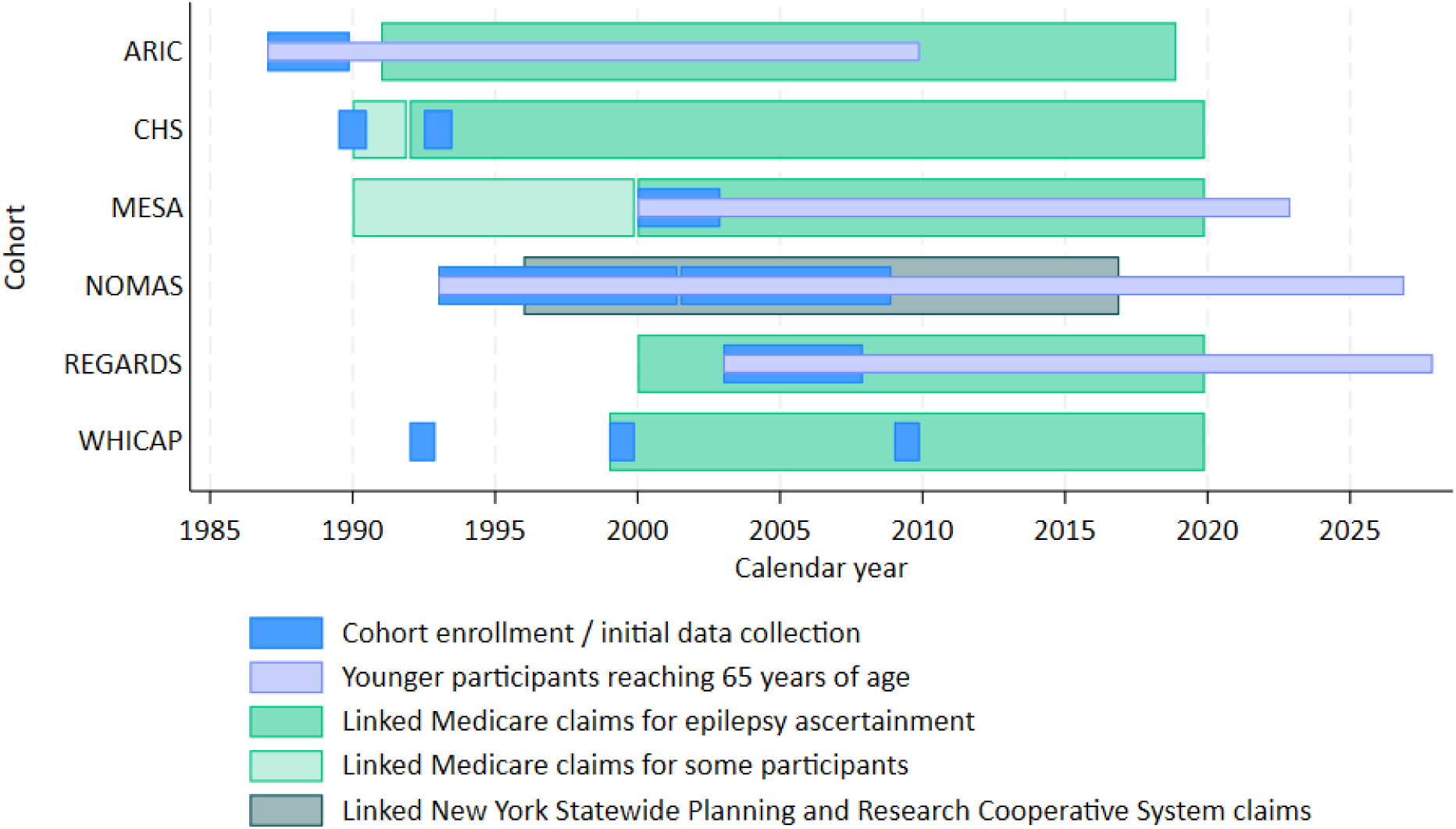
Calendar years of cohort enrollment and availability of linked claims for epilepsy case ascertainment in six cohorts. ARIC, MESA, NOMAS, and REGARDS enrolled participants younger than 65 years of age who then reached 65 years of age during cohort follow-up. CHS and WHICAP enrolled participants 65 years of age and above, so no participants reached 65 years of age during cohort follow-up.

For each study participant, the date on which all eligibility criteria (a), (b), (c), and (d) were met was considered that participant’s “Epilepsy-Cog baseline.” Epilepsy-Cog baseline date could be the date of cohort entry or a later date, depending on each participant’s timeline of Medicare claims availability and continuous Medicare fee-for-service enrollment (Figure 2). Participants in Medicare-linked cohorts were followed for incident epilepsy through the end of their period of continuous enrollment in Medicare fee-for-service. In contrast to the five Medicare-linked cohorts, NOMAS was linked to the New York Statewide Planning and Research Cooperative System (NY-SPARCS) claims, used to capture incident vascular events that may have been missed despite the annual phone interview. However, we did not require NY-SPARCS claims linkage as an eligibility criterion for NOMAS participants in the Epilepsy-Cog study.

**Figure 2.**
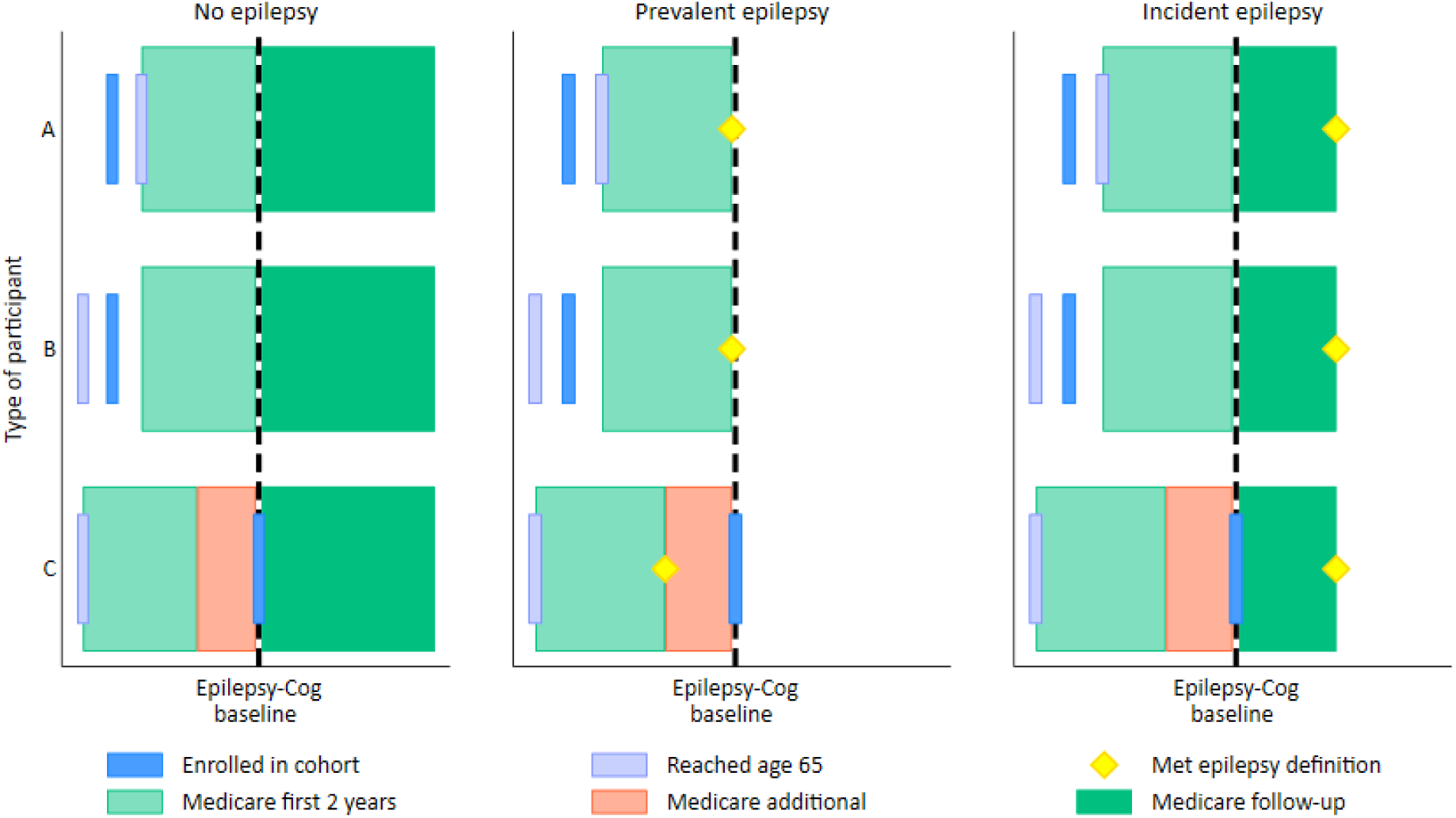
Time alignment of cohort enrollment, aging, and Medicare enrollment to derive Epilepsy-Cog baseline for each participant in five Medicare-linked cohorts. Epilepsy-Cog baseline (bold dashed black vertical line), representing the point at which prevalent epilepsy was determined and follow-up for incident epilepsy began, is shown for participants of three types.

- Type A participants first enrolled in their cohort when younger than 65 years of age, then reached 65 years of age and enrolled in Medicare during cohort follow-up and later reached 2 years of continuous Medicare enrollment. Type A participants were commonly found in ARIC, MESA, and REGARDS. Their Epilepsy-Cog baseline occurred at their Medicare 2-year mark, sometime after cohort enrollment.
- Type B participants first reached 65 years of age (and likely enrolled in Medicare at that time), then enrolled in their cohort when 65 years of age or older, but Medicare claims availability linked to the cohort began after cohort enrollment, thus allowing participants to reach 2 years of continuous Medicare enrollment sometime during cohort follow-up. Type B participants were commonly found in CHS, MESA, REGARDS, and WHICAP. Their Epilepsy-Cog baseline occurred at their Medicare 2-year mark, sometime after cohort enrollment.
- Type C participants first reached 65 years of age and enrolled in Medicare, then enrolled in their cohort when older than 65 years of age, and Medicare claims availability linked to the cohort predated cohort enrollment, thus allowing participants to reach 2 years of continuous Medicare enrollment prior to cohort enrollment. Some Type C participants would have additional time between their Medicare 2-year mark and their cohort enrollment, which provided additional time to ascertain prevalent epilepsy. Type C participants were commonly found in CHS, MESA, REGARDS, and WHICAP. Their Epilepsy-Cog baseline occurred at cohort enrollment.

Claims-based ascertainment of prevalent and incident epilepsy was applicable to participants of all types (A, B, C). Participants who never met a claims-based epilepsy definition would contribute to the denominator for epilepsy prevalence at Epilepsy-Cog baseline and would contribute to the person-time denominator for epilepsy incidence from Epilepsy-Cog baseline until the end of their continuous Medicare enrollment. Participants who met a claims-based definition of epilepsy during the time prior to Epilepsy-Cog baseline (prevalent epilepsy) would contribute to the numerator and denominator for epilepsy prevalence at Epilepsy-Cog baseline and would not contribute any follow-up time for epilepsy incidence after Epilepsy-Cog baseline, since prevalent cases cannot become incident cases. Participants who did not have prevalent epilepsy and who met a claims-based definition of epilepsy during follow-up after Epilepsy-Cog baseline (incident epilepsy) would contribute to the numerator and the person-time denominator for epilepsy incidence from Epilepsy-Cog baseline until the incident epilepsy definition was met.

Rather, we implemented cohort-based epilepsy case ascertainment in NOMAS (see below), so we included all NOMAS participants in Epilepsy-Cog, with Epilepsy-Cog baseline being the date of cohort entry for each NOMAS participant. NOMAS participants were followed for incident epilepsy through the end of the NOMAS cohort follow-up.

### Epilepsy case ascertainment

Epilepsy was not a pre-specified adjudicated outcome in any of the cohort studies. Therefore, we worked with each cohort’s coordinating center to retrospectively ascertain prevalent and incident epilepsy cases.

### Epilepsy case ascertainment in five cohorts linked to Medicare

Data use agreements between the Medicare-linked cohort coordinating centers and the Centers for Medicare and Medicaid Services (CMS) precluded sharing Medicare claims data with investigators outside of the cohort coordinating centers (ARIC: University of North Carolina, Chapel Hill [UNC]; CHS and MESA: University of Washington [UW]; WHICAP: Columbia University; REGARDS: University of Alabama at Birmingham [UAB]). Therefore, for each Medicare-linked cohort, we worked with data managers at the coordinating center to implement Medicare claims-based epilepsy case ascertainment using a standard set of instructions, briefly described here. Among participants we identified as eligible for Epilepsy-Cog (as defined above), we identified epilepsy cases by applying three claims-based epilepsy case definitions that relied on International Classification of Diseases (ICD)-9 or ICD-10 epilepsy diagnoses codes and antiseizure medications, which we adapted from the International League Against Epilepsy’s recommended criteria for classifying epilepsy when using existing coded health data (Supplemental Table 1).^23^ Each of the three case definitions has been previously validated. Case definition 1 has a positive predictive value (PPV), a measure of its ability to identify true cases, ranging from 90 to 99%.^24, 25^ Case definition 2 has a PPV of 91%.^26^ Case definition 3 has a PPV of 84%, 94%, and 98%.^27–29^ The three epilepsy case definitions were: (1) having at least one medical encounter coded with ICD-9 345.xx or ICD-10 G40.x; (2) having at least two medical encounters occurring on different dates, each coded with ICD-9 780.39 or ICD-10 G41x, R56.8, or R56.9; or (3) having at least one medical encounter coded with ICD-9 780.39 or ICD-10 G41x, R56.8, or R56.9, plus at least one occurrence of an antiseizure medication in either (a) Medicare Part D pharmacy claims among those enrolled in Part D (available in some cohorts) or (b) in the cohort medication inventory (available in some cohorts). Each participant could meet any one case definition, any two case definitions, or all three case definitions to qualify as an epilepsy case. Then, epilepsy cases identified by one or more case definitions were further classified as prevalent (pre-existing) epilepsy if at least one of the three case definitions was met prior to Epilepsy-Cog baseline as defined above, or as incident (newly-occurring) LOE if none of the three case definitions were met prior to Epilepsy-Cog baseline and at least one of the three case definitions was met after Epilepsy-cog baseline (Figure 2). If a participant was classified as having prevalent epilepsy by one or more definitions and classified as having incident epilepsy by another definition, the participant was considered to have prevalent epilepsy. Time-to-LOE for incident LOE cases was defined as days elapsed from Epilepsy-Cog baseline to the earliest date of meeting any one of the three epilepsy case definitions after Epilepsy-Cog baseline. Individuals without LOE are followed until the end of continuous Medicare coverage [in A+B-C], due to disenrollment (for more than 1 month) or death.

Four of the Medicare-linked cohorts (ARIC, CHS, MESA, and WHICAP) provided their de-identified derived epilepsy indicators, including prevalent/incident classifications and time-to-LOE variables, to Columbia University and to Brigham Young University (BYU) for analysis.

These data were shared under existing data use agreements between each cohort’s coordinating center and Columbia and, separately, between each center and BYU. The remaining Medicare-linked cohort, REGARDS, is prohibited by its data use agreement with CMS from sharing any Medicare-derived variables, including de-identified derived epilepsy case. Therefore, the REGARDS epilepsy data were not shared with Columbia University or BYU for data analysis but were analyzed separately at UAB.

### Epilepsy case ascertainment in NOMAS

NOMAS participants screened positive for possible epilepsy in three ways: (1) participant responded “yes” to the question “Have you ever had seizures, convulsions or epilepsy?” at cohort entry or during annual telephone follow-up; (2) mention of seizure or epilepsy was identified during a review of hospitalization records at Columbia University Irving Medical Center (where the majority of NOMAS participants receive medical care) or in outside medical records collected when a possible medical event alert was triggered during the annual telephone survey, or (3) one or more ICD-9 codes (345.xx and 780.3x, excluding 345.6x) or ICD-10 codes (G40.xxx and R56.x, excluding G40.82x and R56.0x) were present in the New York Statewide Planning and Research Cooperative System (NY-SPARCS) claims data linked with NOMAS participant data. Once participants with possible epilepsy were identified, a research coordinator reviewed their Columbia University Irving Medical Center electronic medical record to confirm the presence of an epilepsy diagnosis stated in the medical record. If clinical work-up for epilepsy/seizure was mentioned in the medical record but no epilepsy diagnosis was explicitly stated, a NOMAS study neurologist (JG) adjudicated the presence of epilepsy based on all available information. Then, NOMAS epilepsy cases were further classified as prevalent if the condition was present prior to NOMAS cohort entry (prior to the Epilepsy-Cog baseline) or as incident if the condition was not present prior to NOMAS cohort entry and the epilepsy diagnosis occurred during NOMAS follow-up (after the Epilepsy-Cog baseline).

NOMAS’ de-identified epilepsy case indicators, prevalent and incident classification, and time-to-LOE variables are shared between Columbia University and BYU for data analysis under a data use agreement in place between those two institutions.

### Cross-cohort harmonization of sociodemographic characteristics, vascular risk factors, and other variables

#### Stage 1: Data acquisition and preparation

We assembled a core study team, some based at Columbia University and others at BYU, consisting of two neurologists (one specializing in epilepsy and the other in stroke and vascular neurology), an epidemiologist, a biostatistician, two statistical data analysts, and a research coordinator, who met regularly to prepare for and implement data harmonization. This core team consulted as needed with co-investigators involved with each of the six cohorts. We submitted ancillary study proposals to the cohorts’ steering committees, which approved our receipt and use of cohort data. Once the central IRB at Columbia University approved our study and the data use agreements were executed between the data coordinating center of each cohort and Columbia/BYU, the core team reviewed relevant data manuals and documentation from each cohort (such as study protocols, standard operating procedures, and data dictionaries), identified variables to harmonize, and submitted data requests to the cohort coordinating centers. We then received de-identified data from each cohort’s coordinating center. De-identified data from the cohorts and harmonized data are securely maintained on private servers operated by the informatics division of the Department of Neurology at Columbia and by the Department of Public Health at BYU, with access restricted to the core study team. This preparatory stage took place between April 2022 and September 2023.

#### Stage 2: Cross-cohort harmonization

After receiving data from the cohort coordinating centers, we documented every variable by the following data features: (1) original variable name used in a given cohort (e.g., “race”), (2) the construct measured (e.g., race/ethnicity), (3) data type (e.g., categorical), (4) method of data collection (e.g., interview), (5) original response category and value level (e.g., 0=non-Hispanic White, 1=non-Hispanic Black, 2=non-Hispanic Other, and 3=Hispanic), and (6) timing of measurement (e.g., at cohort entry or longitudinally). Grounded in prior work by Levine et al.,^12^ we harmonized variables using the lowest common denominator approach. New variable names and common response categories or value levels were assigned, with variables defined in common across all cohorts, thereby enabling standardized data across all cohorts.

Finally, harmonized datasets for five cohorts (ARIC, CHS, MESA, NOMAS, and WHICAP) were appended with epilepsy case data (described above) and pooled into a single dataset for pooled analysis. As the data use agreement for REGARDS with CMS restricts the sharing of epilepsy variables derived from Medicare outside of UAB, harmonized REGARDS variables were returned to the REGARDS coordinating center at UAB, where they were merged with each participant’s epilepsy data for analysis at UAB. This harmonization stage took place between September 2023 and March 2025.

#### Stage 3: Post-harmonization data validation

Post-harmonization data validation procedures were conducted to confirm that the harmonized data across cohorts were consistent and ready for pooled analyses. This data validation stage took place between March 2025 and April 2025.

### Statistical analysis

We calculated (a) the number of participants meeting eligibility criteria for Epilepsy-Cog pooled cohort among participants of six cohorts (Figure 3), (b) the distribution of time elapsed between cohort entry to Epilepsy-Cog baseline from five Medicare-linked cohorts (Table 2), (c) prevalent and incident epilepsy cases identified among participants across six cohorts (Table 3 and Supplemental Table 2).

**Figure 3.**
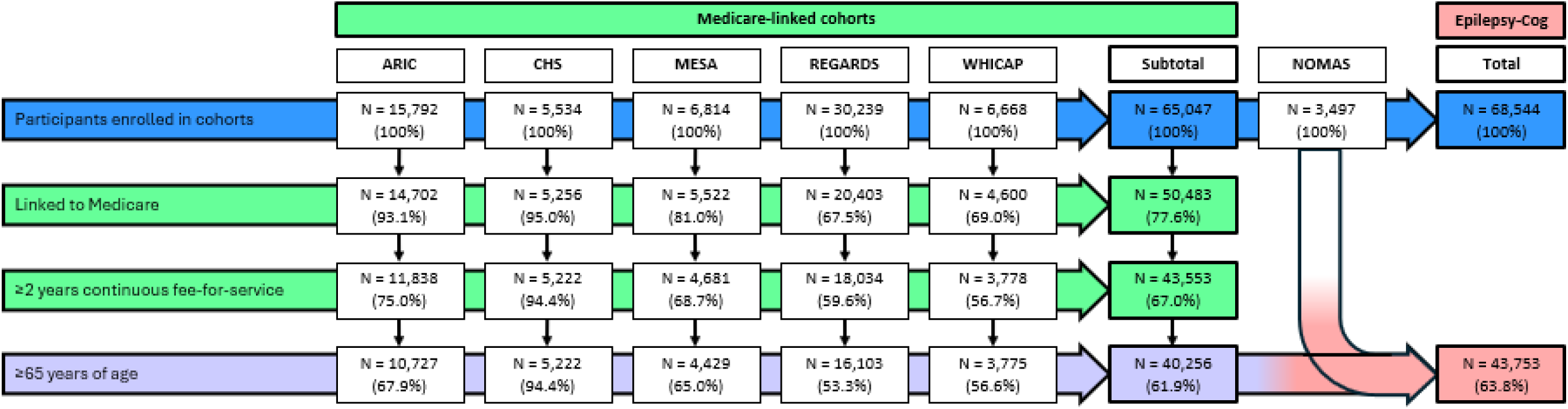
Flowchart of eligibility for Epilepsy-Cog pooled cohort among participants of six cohorts. While CHS enrolled 5,888 participants, we considered only the 5,534 participants who were alive at the 1992/93 exam because CHS Medicare linkage began in 1992 for most participants. While WHICAP enrolled 6,792 participants, we considered only the 6,668 participants who were not also enrolled in NOMAS.

**Table 2.**
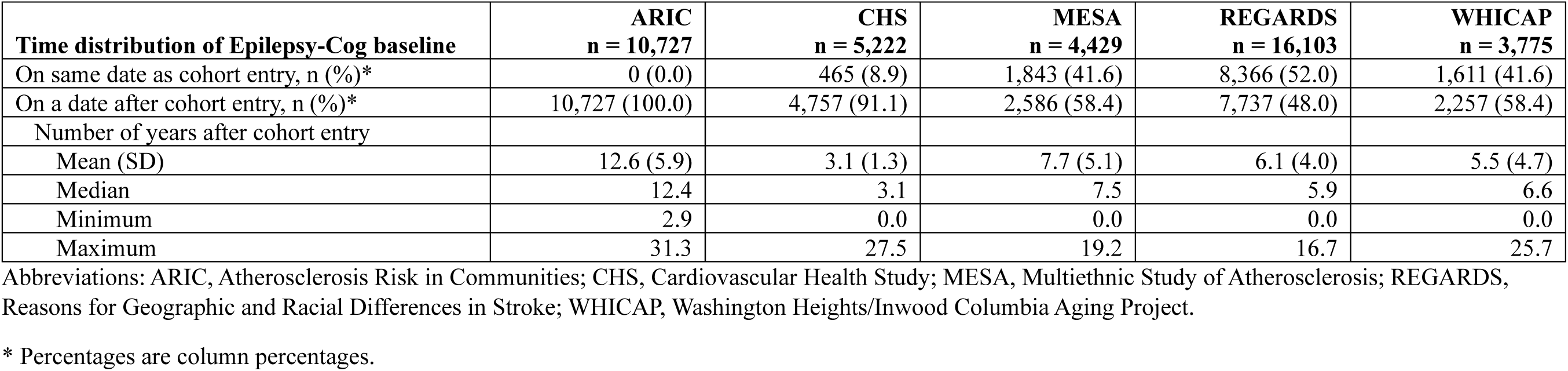
Distribution of time elapsed from cohort entry to Epilepsy-Cog baseline among participants in Epilepsy-Cog from five Medicare-linked cohorts.

**Table 3.**
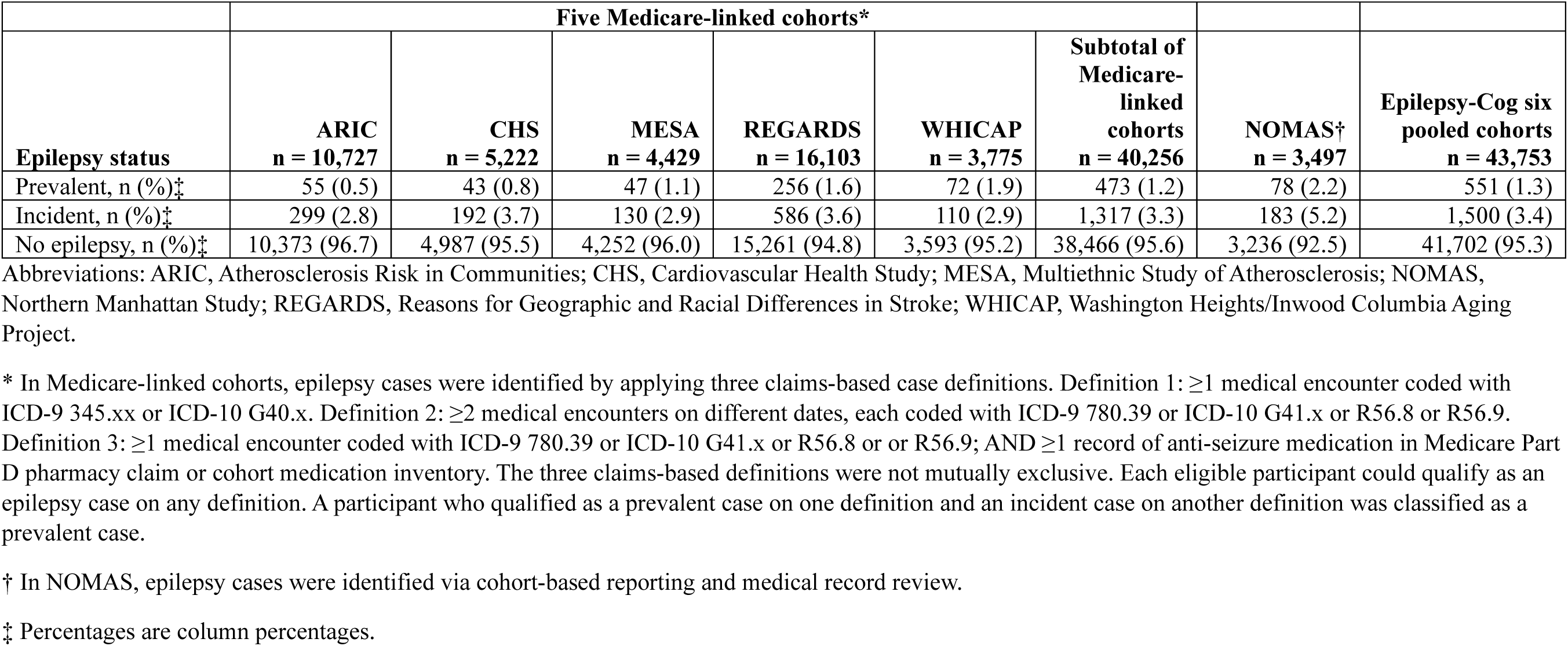
Prevalent and incident epilepsy cases identified among participants in Epilepsy-Cog from six cohorts.

|  | Five Medicare-linked cohorts* |  |  |  |  |  |  |  |
| --- | --- | --- | --- | --- | --- | --- | --- | --- |
| **Epilepsy status** | **ARIC**<br>**n = 10,727** | **CHS**<br>**n = 5,222** | **MESA**<br>**n = 4,429** | **REGARDS**<br>**n = 16,103** | **WHICAP**<br>**n = 3,775** | **Subtotal of**<br>**Medicare-**<br>**linked**<br>**cohorts**<br>**n = 40,256** | **NOMAS†**<br>**n = 3,497** | **Epilepsy-Cog six**<br>**pooled cohorts**<br>**n = 43,753** |
| Prevalent, n (%)‡ | 55 (0.5) | 43 (0.8) | 47 (1.1) | 256 (1.6) | 72 (1.9) | 473 (1.2) | 78 (2.2) | 551 (1.3) |
| Incident, n (%)‡ | 299 (2.8) | 192 (3.7) | 130 (2.9) | 586 (3.6) | 110 (2.9) | 1,317 (3.3) | 183 (5.2) | 1,500 (3.4) |
| No epilepsy, n (%)‡ | 10,373 (96.7) | 4,987 (95.5) | 4,252 (96.0) | 15,261 (94.8) | 3,593 (95.2) | 38,466 (95.6) | 3,236 (92.5) | 41,702 (95.3) |

Abbreviations: ARIC, Atherosclerosis Risk in Communities; CHS, Cardiovascular Health Study; MESA, Multiethnic Study of Atherosclerosis; NOMAS, Northern Manhattan Study; REGARDS, Reasons for Geographic and Racial Differences in Stroke; WHICAP, Washington Heights/Inwood Columbia Aging Project.
\* In Medicare-linked cohorts, epilepsy cases were identified by applying three claims-based case definitions. Definition 1: $\geq 1$ medical encounter coded with ICD-9 345.xx or ICD-10 G40.x. Definition 2: $\geq 2$ medical encounters on different dates, each coded with ICD-9 780.39 or ICD-10 G41.x or R56.8 or R56.9. Definition 3: $\geq 1$ medical encounter coded with ICD-9 780.39 or ICD-10 G41.x or R56.8 or R56.9; AND $\geq 1$ record of anti-seizure medication in Medicare Part D pharmacy claim or cohort medication inventory. The three claims-based definitions were not mutually exclusive. Each eligible participant could qualify as an epilepsy case on any definition. A participant who qualified as a prevalent case on one definition and an incident case on another definition was classified as a prevalent case.
† In NOMAS, epilepsy cases were identified via cohort-based reporting and medical record review.
‡ Percentages are column percentages.

To ensure consistency across two data analysts, we applied data quality control procedures. First, both data analysts harmonized a common set of sociodemographic and VRF variables in three of the six cohorts. Quality assessment was performed by generating and comparing frequency tables for categorical variables and by calculating mean, standard deviation, minimum, maximum, and median values for continuous variables. The analyses were performed using R version 4.5.0 and SAS 9.4 with SAS/STAT 15.2.

## RESULTS

### Medicare linkage and participant eligibility for Epilepsy-Cog pooled cohort

The six cohorts included a total of 68,544 participants (Figure 3). Among the 65,047 participants in the five Medicare-linked cohorts, 50,483 (77.6%) were linked to Medicare, ranging from 67.5% in REGARDS to 95.0% in CHS, consistent with a lower age range of 45+ at cohort entry in REGARDS and 65+ in CHS. Of those who were linked to Medicare, 43,553 (67.0%) had ≥2 years of continuous enrollment in Medicare fee-for-service, ranging from 56.7% in WHICAP to 94.4% in CHS. From those with ≥2 years of continuous Medicare fee-for-service enrollment, we excluded those <65 years of age at Epilepsy-Cog baseline, resulting in 40,256 participants across the five Medicare-linked cohorts. The addition of 3,497 NOMAS participants increased the total Epilepsy-Cog cohort to 43,753 participants (63.8% of participants in the original six cohorts).

### Epilepsy-Cog baseline in Medicare-linked cohorts

Shown in Table 2 are the distributions of time elapsed between cohort entry and Epilepsy-Cog baseline in the five Medicare-linked cohorts. NOMAS is omitted in this analysis since NOMAS used cohort-based epilepsy case ascertainment rather than Medicare-linked case ascertainment.

Because ARIC participants were between the ages of 45 and 64 years at cohort entry, the Epilepsy-Cog baseline for all ARIC participants was at least 2 years after their cohort entry.

Since their age at ARIC entry was younger than the age of Medicare eligibility, they aged into Medicare after ARIC entry, thus their Epilepsy-Cog baseline was a mean of 12.6 years after ARIC entry.

MESA participants were aged 45-84 years at cohort entry. Given this wide age range, some participants aged into Medicare after MESA entry, while others were already in Medicare at MESA entry. Therefore, 58% of the MESA participants had Epilepsy-Cog baseline after MESA entry at a mean of 7.7 years after MESA entry. Similarly, REGARDS participants were aged 45 years and older at cohort entry, and 48% of REGARDS participants had Epilepsy-Cog baseline after REGARDS entry at a mean of 6.1 years after their cohort entry.

CHS and WHICAP participants were aged 65 years or older at cohort entry; therefore, their Epilepsy-Cog baseline would be defined by the availability of linked Medicare claims rather than by aging into Medicare. Epilepsy-Cog baseline was after CHS entry for 91% of CHS participants, with a mean of 3.1 years after CHS cohort entry. Similarly, 58% of WHICAP participants had Epilepsy-Cog baseline after WHICAP cohort entry, at a mean of 5.5 years after WHICAP cohort entry.

### Epilepsy case ascertainment

Shown in Table 3 are counts of prevalent, incident, and non-epilepsy cases in each cohort and in the Epilepsy-Cog pooled cohort of 43,753 participants. The percentage of participants with prevalent epilepsy in the five Medicare-linked cohorts ranged from 0.5% in ARIC to 1.9% in WHICAP. In NOMAS, in which epilepsy cases were identified via cohort-based reporting and medical record review, 2.2% of participants had prevalent epilepsy. In the Epilepsy-Cog pooled cohort, 551 (1.3%) participants had prevalent epilepsy.

The percentage of participants identified as experiencing incident epilepsy in the five Medicare-linked cohorts ranged between 2.8% in ARIC to 3.7% in CHS. In NOMAS, 5.2% of participants had incident epilepsy. In the Epilepsy-Cog pooled cohort, 1,500 (3.4%) participants had incident epilepsy.

Detailed results of applying the three claims-based epilepsy case definitions in the five Medicare-linked cohorts are shown in Supplemental Table 2. Case definition 1, based on ≥1 medical encounter with ICD-9 code 345.xx or ICD-10 code G40.x, yielded the highest prevalence and incident case identification rates compared to Case definitions 2 or 3.

### Cross-cohort harmonization of sociodemographic characteristics, vascular risk factors, and other variables

#### Cross-cohort harmonization

As shown in Supplemental Table 3, we harmonized sociodemographic characteristics: age, sex, race/ethnicity, marital status, educational level, occupation type, and geographic location; health behaviors: smoking and alcohol intake; a variety of VRFs including physical body measures, blood measures, and disease history; APOE genotype; and medications used for conditions including high blood pressure, diabetes, high cholesterol, and antithrombotic treatment. All sociodemographic, health behaviors, vascular risk factors, and medications used for vascular conditions that we harmonized were assessed at cohort entry, and age at Epilepsy-Cog baseline.

Longitudinal subjective health status measures included self-reported health status and depressive symptoms measured with the Center for Epidemiologic Studies Depression (CESD) Scale, assessed at cohort entry and during cohort follow-up. We also harmonized variables for incident events and time-to-event, including incident myocardial infarction, incident atrial fibrillation, incident heart failure, incident stroke, stroke subtypes, and stroke causes. Finally, we harmonized variables related to vital status, including death, time to death, death due to atherosclerotic cardiovascular disease, myocardial infarction, and stroke.

During the harmonization process, we found that the conceptual definitions of the variables we harmonized were entirely consistent across cohorts. In contrast, original coding schemes and values showed incomplete overlap for most categorical variables.

Basic demographic variables were measured similarly across cohorts. We harmonized race/ethnicity using U.S. Census classifications (NH-White, NH-Black, NH-Other, Hispanic, Asian). For example, we recoded White and Black participants in ARIC and REGARDS as NH-White or NH-Black, recognizing that some may have Hispanic ethnicity. We recoded location as residing within or outside the stroke belt.

Across cohorts, smoking status was comparable. We harmonized alcohol intake to drinks per week. Because ARIC measured weekly drink consumption in grams, we divided the weekly intake by 14, the number of grams per drink. We recoded prevalent VRFs at cohort entry (history of hypertension, diabetes, high cholesterol, and chronic kidney disease) using all available data components, including self-report, medication use, and biomarker thresholds.

We harmonized continuous variables, such as body mass index, waist circumference, and blood pressure, by converting units or averaging values. We collapsed APOE genotypes into three groups (ε2 carriers, ε3 homozygotes, ε4 carriers) and also created a three-level APOE ε4 allele status variable.

We harmonized medication use for VRFs at cohort entry using self-reported medication use or medication inventory data available from each cohort. We collapsed subjective health measures into fewer categories and standardized depressive symptoms using a cohort-specific CESD cutoff.

For incident event variables, we collapsed categories when needed to standardize data across cohorts. For example, we collapsed hemorrhagic and ischemic stroke subtype categories to a common form supported by all cohorts.

#### Validation of the cross-cohort harmonization process

All summary statistics values matched between the two data analysts.

## DISCUSSION

We retrospectively harmonized and pooled data from six NIH-funded, US-based, prospective cohort studies of older adults to create the first large-scale, observational, multicohort study of epilepsy, a growing health concern in an aging population. Except in NOMAS, where epilepsy cases were identified through cohort-based reporting and medical record review, we used existing linked Medicare claims data linkage to determine epilepsy, categorizing study participants into those with prevalent, incident, or no epilepsy. This unique effort to create a longitudinal pooled cohort of older adults with and without epilepsy provides a first-ever opportunity to study LOE on a large scale.

Our pooled prevalence estimate of epilepsy of 1.3% in older adults is consistent with prior single-cohort studies despite methodological differences. It closely matches estimates from the Rotterdam study, a population-based door-to-door survey using structured questions on epilepsy and antiseizure medication, which found 1.2% (ages 65-74), 1.1% (ages 75-84), and 1.5% (ages 85-94).^30^ Our estimate is also consistent with U.S. population-based data. The 2021 National Health and Nutrition Examination Survey reported a prevalence of 1.1% among adults aged ≥65 years,^31^ and Medicare claims analyses similarly found an average annual prevalence of 1.1%.^32^

In contrast to prevalence estimates, population-level incidence data with longitudinal follow-up remain relatively scarce.^33^ To date, this Epilepsy-Cog pooled study is the largest of its kind in the U.S.. Estimation of incidence rates with person-time will be included in future analyses. Variation in case definitions between previously published single-cohort studies and our study may help explain the differences in the incidence proportion. Wider adoption of existing case definitions for identifying LOE when using claims-based data will likely address discrepancies in findings.

Across cohorts, the proportion of participants with prevalent or incident epilepsy was higher in NOMAS than in the five Medicare-linked cohorts. This difference may reflect variation in the case ascertainment method. In NOMAS, we first screened for possible epilepsy and subsequently confirmed it through medical record review, given its lack of linkage to Medicare claims. Consequently, misclassification of epilepsy cases may have differed between NOMAS and those of the other five cohorts. Additionally, differences in estimates between NOMAS and other cohorts may reflect underlying population differences. For example, nearly two-thirds of NOMAS participants were Hispanic or non-Hispanic Black. Racial differences in LOE incidence have been previously reported.^6, 32, 34^

The benefits of pooling individual participant data from multiple cohorts through data harmonization are many.^35^ First, it is less costly and more time-efficient to utilize existing longitudinal data than to collect primary epilepsy data in a new study and follow study participants for an extended period. Our Epilepsy-Cog pooled cohort study includes 43,753 participants; designing a prospective, longitudinal study of similar magnitude to examine the consequences of LOE would be cost-prohibitive. Second, in contrast to prior single-population-based studies of LOE, whose sample sizes ranged from 55 to 678 cases with LOE, our pooled sample of 1,500 LOE cases would yield lower standard errors and greater statistical power to detect smaller effect sizes. Third, by combining individual-level data from multiple studies through data harmonization, the issue of data heterogeneity across different cohorts can be addressed more easily than in a meta-analysis of aggregated data.

Nevertheless, the data harmonization process presented several challenges. First, it required a series of rigorous and complex steps, including developing and implementing the epilepsy case ascertainment algorithm, identifying variables for harmonization, determining how each original variable was assessed across cohorts, establishing harmonization decision rules, implementing those rules, and conducting quality reviews. Second, because harmonization relied on existing data, the extent of missingness varied across studies. In future analyses, we will employ strategies to mitigate potential bias arising from missing data. Third, although consensus guidelines on best practices for data harmonization were available,^36^ some challenges noted in the guidelines were unavoidable in our context, particularly the time needed to access cohort data and to learn cohort-specific designs and standard operating procedures.

The purpose of creating this pooled cohort is to eventually investigate relationships of LOE, pre-epilepsy VRFs, with post-LOE stroke and cognitive decline. Epilepsy-Cog has three specific aims we hope to accomplish: (1) Quantify the association of LOE with cognitive decline, and identify the roles of pre-epilepsy VRFs and MRI-defined covert vascular brain injury in this association; (2) Assess whether LOE, as a novel risk marker of covert vascular brain injury, changes predicted risks for Alzheimer’s disease and stroke when incorporated into established risk prediction algorithms; (3) Use microsimulation to quantify the incremental value of risk/benefit-based tailored treatment of VRFs, compared with traditional treat-to-target approaches, on reducing Alzheimer’s disease and stroke risk after LOE. As no single cohort will be large enough to pursue these aims, Epilepsy-Cog is anticipated to advance understanding of LOE.

## Conclusion

We retrospectively pooled and harmonized data from six NIH-funded, US-based, prospective cohort studies of cardiovascular, stroke, and cognitive outcomes in older adults and leveraged their linked Medicare data to ascertain epilepsy. With 551 prevalent epilepsy and 1,500 incident epilepsy cases identified from our Epilepsy-Cog pooled cohort of 43,753 participants, combined with harmonized demographic, vascular risk factors, and events data, this pooled study presents an unprecedented opportunity to understand factors contributing to the development of LOE and the risks for the development of subsequent adverse events following LOE in older age.

## Funding Sources

**<u>Epilepsy-Cog</u>**: This work was supported by the National Institutes of Health (grant R01AG074355). Drs. Thacker, Choi, and Gutierrez were supported by this grant.

**<u>ARIC</u>**: The Atherosclerosis Risk in Communities study has been funded in whole or in part with Federal funds from the National Heart, Lung, and Blood Institute, National Institutes of Health, Department of Health and Human Services, under Contract nos. (75N92022D00001, 75N923022D00002, 75N92022D00003, 75N92022D00004, 75N92022D00005). The authors thank the staff and participants of the ARIC study for their important contributions.

**<u>CHS</u>**: This research was supported by contracts HHSN268201200036C, HHSN268200800007C, HHSN268201800001C, N01HC55222, N01HC85079, N01HC85080, N01HC85081, N01HC85082, N01HC85083, N01HC85086, 75N92021D00006, and grants U01HL080295, U01HL130114, and R01HL172803 from the National Heart, Lung, and Blood Institute (NHLBI), with additional contribution from the National Institute of Neurological Disorders and Stroke (NINDS). Additional support was provided by R01AG023629 from the National Institute on Aging (NIA). A full list of principal CHS investigators and institutions can be found at CHS-NHLBI.org.

**MESA:** MESA is supported by contracts 75N92025D00022, 75N92020D00001, HHSN268201500003I, N01-HC-95159, 75N92025D00026, 75N92020D00005, N01-HC-95160, 75N92020D00002, N01-HC-95161, 75N92025D00024, 75N92020D00003, N01-HC-95162, 75N92025D00027, 75N92020D00006, N01-HC-95163, 75N92025D00025, 75N92020D00004, N01-HC-95164, 75N92025D00028, 75N92020D00007, N01-HC-95165, N01-HC-95166, N01-HC-95167, N01-HC-95168 and N01-HC-95169 from the National Heart, Lung, and Blood Institute, and by grants UL1-TR-000040, UL1-TR-001079, and UL1-TR-001420 from the National Center for Advancing Translational Sciences (NCATS).

**NOMAS:** This project has been funded at least in part with federal funds from the National Institutes of Health, National Institute of Neurological Disorders and Stroke by R01-NS29993.

**<u>REGARDS:</u>** This research project is supported by cooperative agreement U01 NS041588 co-funded by the National Institute of Neurological Disorders and Stroke (NINDS) and the National Institute on Aging (NIA), National Institutes of Health, Department of Health and Human Service. The content is solely the responsibility of the authors and does not necessarily represent the official views of the NINDS or the NIA. Representatives of the NINDS were involved in the review of the manuscript but were not directly involved in the collection, management, analysis or interpretation of the data. The authors thank the other investigators, the staff, and the participants of the REGARDS study for their valuable contributions. A full list of participating REGARDS investigators and institutions can be found at: https://www.uab.edu/soph/regardsstudy/.

**<u>WHICAP</u>**: Data collection and sharing for this project was supported by the Washington Heights-Inwood Columbia Aging Project (WHICAP,R01AG072474, RF1AG066107, PO1AG07232, R01AG037212, RF1AG054023) funded by the National Institute on Aging (NIA). This manuscript has been reviewed by WHICAP investigators for scientific content and consistency of data interpretation with previous WHICAP Study publications. We acknowledge the WHICAP study participants and the WHICAP research and support staff for their contributions to this study

## Conflict of Interest Statement

The authors declare no conflicts of interest.

## Data Availability

De-identified participant-level data from the cohort studies included in this analysis are available through the respective coordinating center of each cohort, subject to study-specific data access policies and approvals. Information on data access procedures is publicly available online.

https://sites.cscc.unc.edu/aric

https://chs-nhlbi.org

https://mesa-nhlbi.org

https://med.miami.edu/departments/neurology/research/nomas/nomas-policies

https://uab.edu/soph/regardsstudy/researchers

https://columbianeuroresearch.org/taub/res-normal.html

---

**Supplemental Table 1.** Claims-based epilepsy case definitions adapted from the International League Against Epilepsy recommended criteria.

| **Definition** | **Qualifying medical encounters** | **Qualifying ICD codes** | **Anti-seizure medications** |
| --- | --- | --- | --- |
| Definition 1 | ≥1 medical encounter occurring on a date between the beginning and end of the participant's continuous enrollment in Medicare A+B-C±D with a claim containing one of the qualifying codes in any position | ICD-9 345.xx; ICD-10 G40.x | Not used in definition 1 |
| Definition 2 | ≥2 medical encounters occurring on different dates, both between the beginning and end of the participant's continuous enrollment in Medicare A+B-C±D, with each claim containing at least one of the qualifying codes in any position | ICD-9 780.39; ICD-10 G41.x or R56.8 or R56.9 | Not used in definition 2 |
| Definition 3 | ≥1 medical encounter occurring on a date between the beginning and end of the participant's continuous enrollment in Medicare A+B-C±D with a claim containing one of the qualifying codes in any position | ICD-9 780.39; ICD-10 G41.x or R56.8 or R56.9 | AND ≥1 record of AED, which may be either of the following: (1) pharmacy claim in Medicare Part D containing a code for any anti-seizure medication; (2) cohort medication inventory containing any anti-seizure medication |

**Supplemental Table 2.**
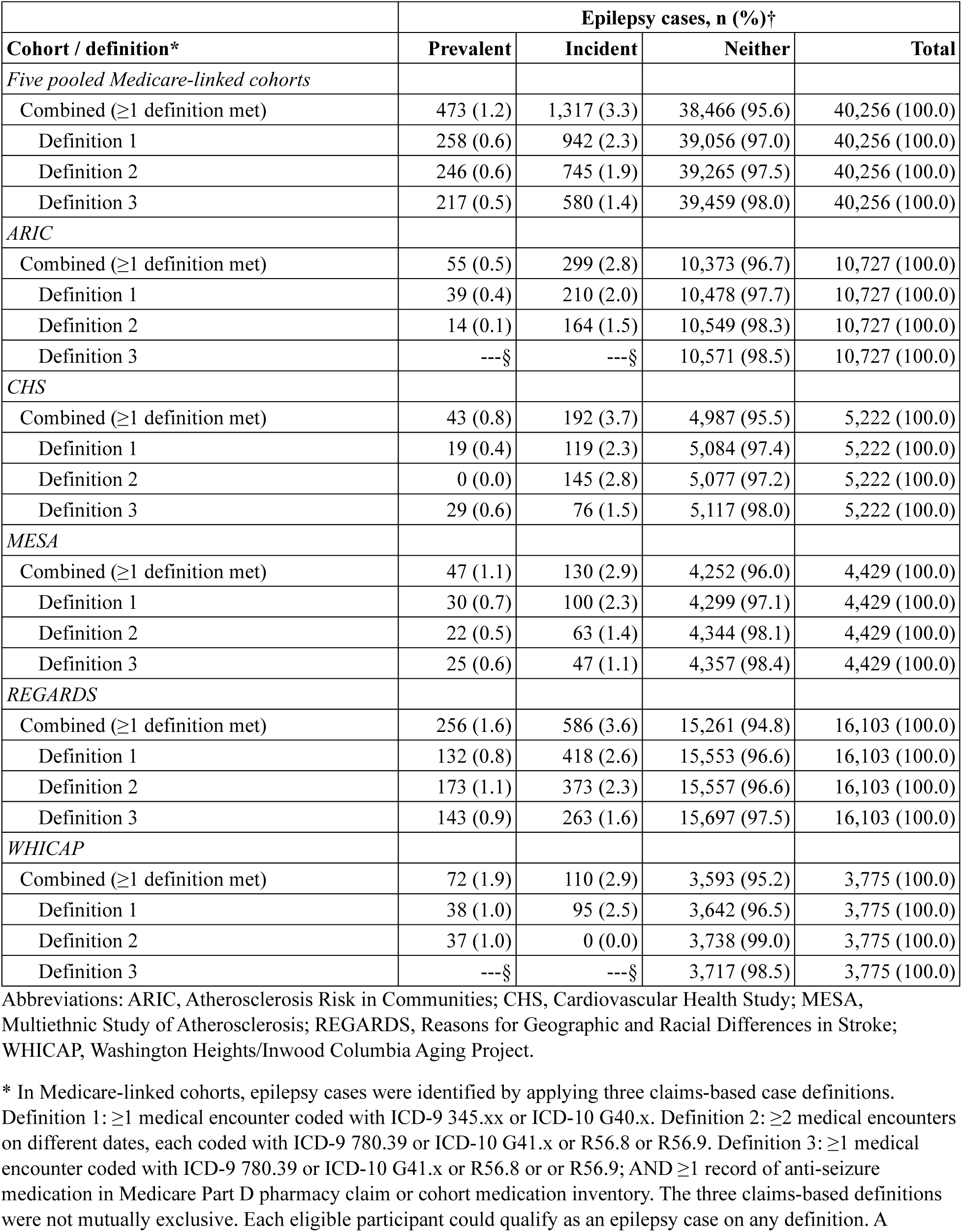
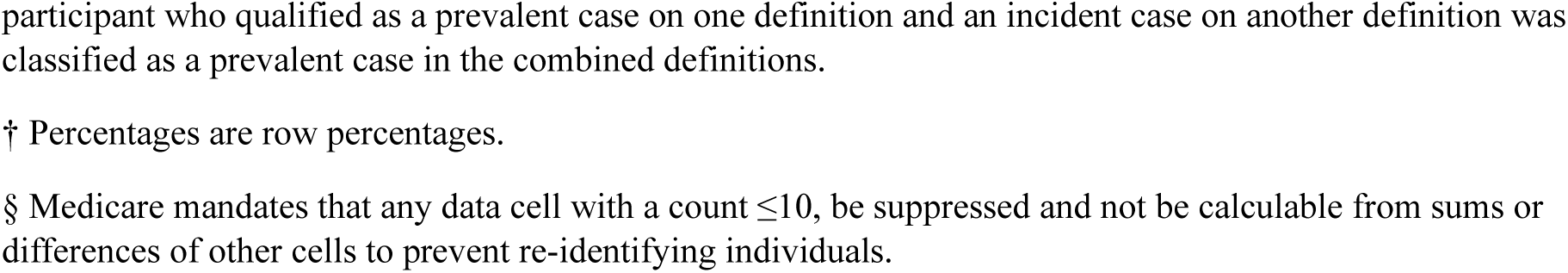
Prevalent and incident epilepsy cases identified by three claims-based definitions among Epilepsy-Cog participants from five Medicare-linked cohorts.

| Cohort / definition* | Epilepsy cases, n (%)† |  |  |  |
| --- | --- | --- | --- | --- |
|  | Prevalent | Incident | Neither | Total |
| *Five pooled Medicare-linked cohorts* |  |  |  |  |
| Combined (≥1 definition met) | 473 (1.2) | 1,317 (3.3) | 38,466 (95.6) | 40,256 (100.0) |
| Definition 1 | 258 (0.6) | 942 (2.3) | 39,056 (97.0) | 40,256 (100.0) |
| Definition 2 | 246 (0.6) | 745 (1.9) | 39,265 (97.5) | 40,256 (100.0) |
| Definition 3 | 217 (0.5) | 580 (1.4) | 39,459 (98.0) | 40,256 (100.0) |
| *ARIC* |  |  |  |  |
| Combined (≥1 definition met) | 55 (0.5) | 299 (2.8) | 10,373 (96.7) | 10,727 (100.0) |
| Definition 1 | 39 (0.4) | 210 (2.0) | 10,478 (97.7) | 10,727 (100.0) |
| Definition 2 | 14 (0.1) | 164 (1.5) | 10,549 (98.3) | 10,727 (100.0) |
| Definition 3 | ---§ | ---§ | 10,571 (98.5) | 10,727 (100.0) |
| *CHS* |  |  |  |  |
| Combined (≥1 definition met) | 43 (0.8) | 192 (3.7) | 4,987 (95.5) | 5,222 (100.0) |
| Definition 1 | 19 (0.4) | 119 (2.3) | 5,084 (97.4) | 5,222 (100.0) |
| Definition 2 | 0 (0.0) | 145 (2.8) | 5,077 (97.2) | 5,222 (100.0) |
| Definition 3 | 29 (0.6) | 76 (1.5) | 5,117 (98.0) | 5,222 (100.0) |
| *MESA* |  |  |  |  |
| Combined (≥1 definition met) | 47 (1.1) | 130 (2.9) | 4,252 (96.0) | 4,429 (100.0) |
| Definition 1 | 30 (0.7) | 100 (2.3) | 4,299 (97.1) | 4,429 (100.0) |
| Definition 2 | 22 (0.5) | 63 (1.4) | 4,344 (98.1) | 4,429 (100.0) |
| Definition 3 | 25 (0.6) | 47 (1.1) | 4,357 (98.4) | 4,429 (100.0) |
| *REGARDS* |  |  |  |  |
| Combined (≥1 definition met) | 256 (1.6) | 586 (3.6) | 15,261 (94.8) | 16,103 (100.0) |
| Definition 1 | 132 (0.8) | 418 (2.6) | 15,553 (96.6) | 16,103 (100.0) |
| Definition 2 | 173 (1.1) | 373 (2.3) | 15,557 (96.6) | 16,103 (100.0) |
| Definition 3 | 143 (0.9) | 263 (1.6) | 15,697 (97.5) | 16,103 (100.0) |
| *WHICAP* |  |  |  |  |
| Combined (≥1 definition met) | 72 (1.9) | 110 (2.9) | 3,593 (95.2) | 3,775 (100.0) |
| Definition 1 | 38 (1.0) | 95 (2.5) | 3,642 (96.5) | 3,775 (100.0) |
| Definition 2 | 37 (1.0) | 0 (0.0) | 3,738 (99.0) | 3,775 (100.0) |
| Definition 3 | ---§ | ---§ | 3,717 (98.5) | 3,775 (100.0) |

Abbreviations: ARIC, Atherosclerosis Risk in Communities; CHS, Cardiovascular Health Study; MESA, Multiethnic Study of Atherosclerosis; REGARDS, Reasons for Geographic and Racial Differences in Stroke; WHICAP, Washington Heights/Inwood Columbia Aging Project.
\* In Medicare-linked cohorts, epilepsy cases were identified by applying three claims-based case definitions. Definition 1: ≥1 medical encounter coded with ICD-9 345.xx or ICD-10 G40.x. Definition 2: ≥2 medical encounters on different dates, each coded with ICD-9 780.39 or ICD-10 G41.x or R56.8 or R56.9. Definition 3: ≥1 medical encounter coded with ICD-9 780.39 or ICD-10 G41.x or R56.8 or R56.9; AND ≥1 record of anti-seizure medication in Medicare Part D pharmacy claim or cohort medication inventory. The three claims-based definitions were not mutually exclusive. Each eligible participant could qualify as an epilepsy case on any definition. A
participant who qualified as a prevalent case on one definition and an incident case on another definition was classified as a prevalent case in the combined definitions.
† Percentages are row percentages.
§ Medicare mandates that any data cell with a count $\leq 10$ , be suppressed and not be calculable from sums or differences of other cells to prevent re-identifying individuals.

**Supplemental Table 3.** Harmonization of sociodemographic characteristics, health behaviors, vascular risk factors, incident vascular events, genetic factors, medication use, and causes of death. Harmonization of cognitive measures is a part of this study but will be presented in future papers.

| Type | Variables | Harmonization process | Variables combined and recoded |
| --- | --- | --- | --- |
|  | Participant ID | Action taken: Recoding by concatenating original study ID and study name | Concatenated original study ID and study name |
| **Sociodemographic characteristics** |  |  |  |
|  | Age | Action taken: Renamed the variable to be consistent across all cohorts | Variables renamed, age at cohort entry, and also age at Epilepsy-Cog baseline |
|  | Sex | Complete matching in coding schemes and value levels. Action taken: Variables recoded from male/female to Female, yes/no. | Variables were recoded as Female, Yes/No |
|  | Race/ethnicity | Partial matching in coding schemes and value levels. Actions taken: the original cohort-specific variable was recoded into a 5-category race/ethnicity variable. This was later collapsed into 4-category and 2-category versions for analysis. ARIC did not collect information on ethnicity. In the REGARDS cohort, only Black and White categories were available; all other race/ethnicity values were coded as 0 in those cases. | Combined and recoded to create 3 different variables:<br>Race/ethnicity 5 category (Non-Hispanic white, Non-Hispanic black, Non-Hispanic other, Hispanic, Asian)<br>Race/ethnicity 4 category (Non-Hispanic white, Non-Hispanic black, Non-Hispanic Asian or other, Hispanic)<br>Race/ethnicity 2 category (Non-Hispanic white, All other) |
|  | Marital status | Partial matching in coding schemes and value levels. Action taken: We combined and recoded variables to create marital status 5 category at cohort entry. This was later collapsed into 2-category version for analysis. | Variables were combined and recoded as:<br>Marital status 5 category at cohort entry (Married/living as married, Widowed, Divorced, Separated, Never married/single)<br>Marital status 2 category at cohort entry (Married/living as married, Widowed/divorced/separated/never married/single) |
|  | Education | Partial matching in coding schemes and value levels. Action taken: education categories were combined and recoded into 4 categories in all cohorts except REGARDS, where education data aligned with the new variable categories. | Variables were combined and recoded as follows:<br>Response Categories:<br>1 Less than high school graduate<br>2 High school grad/GED<br>3 Some college but no degree<br>4 College graduate or more |
|  | Occupation type | Partial matching construct (longest or current occupation). In four cohorts (ARIC, CHS, NOMAS, REGARDS, and WHICAP), the longest-held occupation was assessed. MESA assessed the current occupation. | Harmonized longest-held occupation type into four categories:<br>1 White collar<br>2 Blue collar<br>3 Homemaker<br>4 other. |
|  | Geographic location | Action taken: Original study site recoded into a new variable Stroke Belt. Stroke Belt defined as the states of Alabama, Arkansas, Louisiana, Mississippi, Tennessee and the noncoastal regions within the states of North Carolina, South Carolina, and Georgia) and the stroke buckle (defined as the coastal regions within the states of North Carolina, South Carolina, and Georgia). | Variables were combined and recoded as Stroke belt yes/no |
| **Health behaviors** |  |  |  |
|  | Smoking | Partial matching in coding schemes and value levels. Action taken: ARIC, CHS, REGARDS, NOMAS, and MESA had complete category matches. In WHICAP we combined and recoded variables including current smoking status, history of smoking at least one cigarette per day for a year, and age started/stopped smoking. | Combined and recoded as follows:<br>0=Never<br>1=Former<br>2=Current |
|  | Alcohol | Partial matching in coding schemes and value levels. Action taken: In ARIC, usual ethanol intake (grams/week) was converted to drinks/week by dividing by 14 (1 drink = 14g alcohol).<br>*\*References: NutritionHeart, European Heart Journal.* In all other cohorts, drinks/week was recoded if reported in a different time unit. Data not available in WHICAP. | Recoded and combined to create two new variables:<br>1. Drinks per week (Numerical value)<br>2. Drinks per week category:<br>0=None, 1=>0 & <=6, 2=>6 & <=13 ,<br>3=>13 |
| **Vascular risk factors** |  |  |  |
|  | Waist circumference | Partial matching of the unit of measurement.<br>Action taken: Recoding of NOMAS variable by converting inches to centimeters. | Variable converted to measurement in cm for one cohort (waist in cm) |
|  | Waist to hip ratio | Partial matching of the unit of measurement.<br>Action taken: Converted NOMAS hip measurement variable to cm and calculated waist-to-hip ratio by using both hip and waist circumference. For CHS, MESA and WHICAP, calculated waist-to-hip ratio based on their waist and hip circumference. | Variables combined and converted into waist to hip ratio in cm (waist-hip ratio in cm) |
|  | Body mass index | Action taken: BMI (kg/m<sup>2</sup>) from each cohort was recoded to create 3- and 4-category variables. | 3 variables created:<br>Variable coded as BMI (kg/m<sup>2</sup>) at cohort entry<br>BMI 4 category at cohort entry (Underweight, Normal weight, Overweight, Obesity)<br>BMI 3 category at cohort entry (Normal weight or below, Overweight, Obesity) |
|  | Systolic blood pressure | Partial matching in coding.<br>Action taken: In ARIC, CHS, NOMAS, REGARDS, and WHICAP we used SBP measurements as provided (already averaged). For MESA, SBP was averaged from the 2nd and 3rd readings. | Created new SBP variable that calculated the average of readings available, 2 or 3 |
|  | Diastolic blood pressure | Action taken:<br>In ARIC, CHS, NOMAS, REGARDS, and WHICAP we used DBP measurements as provided (already averaged). For MESA, DBP was averaged from the 2nd and 3rd readings. | Created new DBP variable that calculated the average of readings available, 2 or 3 |
|  | Hemoglobin A1C | Did not harmonize; not available in ARIC, CHS, MESA, NOMAS | hba1c (%) |
| | Fasting glucose | Action taken: We recoded the fasting blood draw variable. A fasting indicator was available in ARIC, NOMAS, REGARDS, and MESA. In CHS, fasting status was determined using "hours since last eaten." WHICAP lacked fasting data, so samples were assumed non-fasting. To define diabetes, we used the following rule: fasting glucose $\geq 126$ mg/dL or non-fasting glucose $\geq 200$ mg/dL. | Created 2 variables:<br>Glucose mg/dL at cohort entry<br>Participant was fasting at blood draw (yes/no) |
|  | Total cholesterol | Complete matching unit. | Total cholesterol mg/dL at cohort entry |
|  | LDL cholesterol | Complete matching unit | LDL cholesterol mg/dL at cohort entry |
|  | HDL cholesterol | Complete matching unit | HDL cholesterol mg/dL at cohort entry |
|  | Triglycerides | Complete matching unit | Triglyceride mg/dL at cohort entry |
|  | C-reactive protein | Complete unit. Not available in ARIC, WHICAP. | C-reactive protein mg/L at cohort entry |
|  | Interleukin-6 | Complete matching unit. Not available in ARIC, WHICAP, or REGARDS. | Interleukin-6 pg/ml at cohort entry |
| | History of chronic kidney disease | Partial matching in coding schemes and value levels. Variables created based on patient self-report during study interview and calculated eGFR at cohort entry for each cohort except: Self report was not available in ARIC, NOMAS, or WHICAP. We calculated eGFR as follows: calculate eGFR (estimated glomerular filtration rate) as a single equation.<br>$eGFR_{cr} = 142 * \min(Scr/\kappa, 1)^\alpha * \max(Scr/\kappa, 1) - 1.200 * 0.9938Age * 1.012 \text{ [if female]}$<br>where:<br>Scr = standardized serum creatinine in mg/dL<br>$\kappa = 0.7$ (females) or $0.9$ (males)<br>$\alpha = -0.241$ (female) or $-0.302$ (male)<br>$\min(Scr/\kappa, 1)$ is the minimum of $Scr/\kappa$ or $1.0$<br>$\max(Scr/\kappa, 1)$ is the maximum of $Scr/\kappa$ or $1.0$<br>Age (years)<br>If $eGFR < 60$, it is abnormal or “1”. If $eGFR$ is 60 or higher, then “0”<br>History of chronic kidney disease was considered "missing" if self-report variable was missing along with biomarker for eGFR calculation. | Created 3 variables:<br>Chronic kidney disease at cohort entry (yes/no)<br>Creatinine $eGFR < 60$ at cohort entry (yes/no)<br>NEW chronic kidney disease at cohort entry (yes/no) Created by combining 2 of the variables above |
|  | History of hypertension | Partial matching in coding schemes and value levels. Action taken: This variable was created using three components: self-reported hypertension history, medication use at interview, and blood pressure measured at the study visit. Hypertension was defined by the presence of any one of these, depending on available data from each cohort. Individuals with average SBP $\geq 140$ mmHg or DBP $\geq 90$ mmHg were classified as hypertensive. | Combined and recoded: Hypertension at cohort entry (yes/no) |
| | History of diabetes mellitus | Partial matching in coding schemes and value levels. Action taken: We used the following three components from each cohort (at least one was needed) to create new binary variable: self-report of history of diabetes, medication use, and fasting glucose measurement.<br>We considered fasting glucose $\geq 126$ mg/dL or non-fasting (random) glucose $\geq 200$ mg/dL or Hemoglobin A1c $\geq 6.5\%$ (if available) | Combined and recoded: Diabetes at cohort entry (yes/no) |
|  | History of high cholesterol | Partial matching in coding schemes and value levels. Action taken: a new binary variable was created for the history of high cholesterol. Hyperlipidemia was determined using three components (at least one was needed): self-reported history, medication use, and measured cholesterol levels at cohort entry. | Combined and recoded: Hypercholesterolemia at cohort entry (yes/no) |
|  | History of coronary heart disease/coronary artery disease | Complete matching coding schemes and value levels. Action taken: Coded as "No" for all MESA participants (exclusion criterion). In ARIC, CHS, REGARDS, and NOMAS, the new variable was derived from available self-report, procedure history, and/or ECG data.<br>Data not available in WHICAP. | Combined and recoded: Coronary heart disease at cohort entry (yes/no) |
|  | History of myocardial infarction/heart attack | Partial matching in coding schemes and value levels. Action taken: This variable was coded as "No" for all MESA participants, as the condition was an exclusion criterion. For all other cohorts, we used all available data—such as self-report, ECG-determined MI at cohort entry, and medical history—to recode the variable. | Combined and recoded: History of myocardial infarction at cohort entry (yes/no) |
|  | History of atrial fibrillation | Complete matching coding schemes and value levels. Action taken: This variable was created using all available sources of information across cohorts, including self-report and cohort entry ECG findings. Notably, while MESA excluded participants with known atrial fibrillation (AFib), some cases were identified at cohort entry by electrocardiogram. WHICAP did not have atrial fibrillation data available. | Combined and recoded: Atrial fibrillation at cohort entry (yes/no) |
|  | History of heart failure | Partial matching coding schemes and value levels. Action taken: new variable set to "No" in MESA; data not available in REGARDS. The new HF variable was derived from self-report, medication, or adjudication in other cohorts. We combined and recoded cardiac disease at cohort entry variable and coded as 1 | Two variables created: Heart failure at cohort entry (yes/no)<br>Cardiac disease at cohort entry (MI or CHD or AF or HF) (yes/no) |
|  |  | (yes) if participant had any 1 or more of MI, CHD, AF, HF; 0 (no) if participant had none of those; and missing if criteria for 1 or 0 not met. |  |
|  | History of stroke | Complete matching coding schemes, value levels. Action taken: For MESA and NOMAS, this variable was coded as "No" for all participants, as the condition was an exclusion criterion in both studies. In all other cohorts, we used all available data including self-report or adjudicated medical history. | Combined and recoded: History of stroke at cohort entry (yes/no) |
| **Genetic factor** |  |  |  |
|  | Apolipoprotein E (APOE) Genotype | Partial matching in coding scheme and value levels. Action taken: We used APOE genotype data from each cohort to create three variables. Genotypes were first standardized into six categories: 22, 23, 24, 33, 34, 44. From this, we derived a 3-category variable (E2 carriers, E3 homozygotes, E4 carriers) and a variable indicating the number of APOE ε4 alleles (0, 1, or 2). Data required to construct the 6- and 3-category variables was not available in MESA. | Three variables created:<br>APOE genotype 6 category (e2e2, e2e3, e2e4, e3e3, e3e4, e4e4)<br>APOE genotype 3 category (e2e2/e2e3, e3e3, e2e4/e3e4/e4e4)<br>Number of APOE e4 alleles (0 e4 alleles, 1 e4 allele, 2 e4 alleles) |
| **Medication use** |  |  |  |
|  | Treatment for high BP | Partial matching in coding scheme. Action taken: Self-reported current treatment for high blood pressure or medication inventory from each cohort was used to recode and create a binary variable. | Combined and recoded: Medication for blood pressure at cohort entry (yes/no) |
|  | Treatment for diabetes (insulin or other anti-diabetic meds) | Partial matching in coding scheme. Action taken: We used self-reported current medication use (insulin, oral hypoglycemics, or other anti-diabetic medications) or the medication inventory to recode and create a binary variable for diabetes treatment. | Combined and recoded: Medication for diabetes at cohort entry (yes/no) |
|  | Treatment for high cholesterol | Partial matching in coding scheme. Action taken: We created a binary variable for current high cholesterol treatment using self-reported statin or medication inventory from each cohort. | Combined and recoded: Medication for cholesterol at cohort entry (yes/no) |
|  | Antithrombotic treatment for CVD | Partial matching in coding scheme. Action taken: We created a binary variable for current antithrombotic treatment by recoding self-reported use or medication inventory of aspirin, anticoagulants, or other antiplatelet medications from each cohort. | Combined and recoded: Medication for antithrombosis at cohort entry (yes/no) |
| **Subjective health status (longitudinal)** |  |  |  |
|  | Center for Epidemiologic Studies Depression Scale | Partial matching coding. Action taken: We used cohort-specific cutoff scores in CHS, NOMAS, MESA, and WHICAP to create a binary depression variable. Data not available in ARIC (not assessed at cohort entry) or REGARDS (used a 4-item CES-D). | Combined and recoded: Depression (CESD binary) at cohort entry and during cohort follow-up (yes/no) |
|  | Self-reported health status | Complete matching coding. Action taken: The response coding in MESA was reversed and was recoded to align with other cohorts. The original 5-category variable was then collapsed into 3 categories. Self-reported health status was not available in ARIC (not asked at cohort entry) or WHICAP. | Created two variables;<br>Self-rated health 5 category at cohort entry and during cohort follow-up (Excellent, Very good, Good, Fair, Poor)<br>Self-rated health 3 category at cohort entry and during cohort follow-up (Excellent/very good, Good, Fair/poor) |
| **Incident vascular events** |  |  |  |
|  | Incident MI (definite/probable) | Partial matching coding scheme and value levels. Action taken: We recoded the original cohort-adjudicated variable to include only definite and probable new (incident) cases, excluding anyone who had the event before joining the study. In MESA, MI was an exclusion criterion, so no prior (prevalent) cases were expected. WHICAP did not have data on new MI cases. | Combined and recoded: Incident myocardial infarction yes/no |
|  | Follow-up time to incident MI | Action taken: Follow-up time from cohort entry was standardized to days across cohorts; units were converted when needed. This variable was not available in WHICAP. | Combined and recoded: Days to incident myocardial infarction |
|  | Incident atrial fibrillation | Complete matching coding scheme and value levels. Action taken: We used adjudicated data from all cohorts (except WHICAP and REGARDS, for which data were unavailable) to create a new binary variable. | Combined and recoded: Incident atrial fibrillation (yes/no) |
|  | Follow-up time to incident atrial fibrillation | Complete matching coding scheme. Action taken: in NOMAS, the follow-up interview date when AFib was reported was used to calculate days from cohort entry. Data not available in WHICAP or REGARDS. Because the time windows between visits in REGARDS is high, did not use this data from REGARDS. | Combined and recoded: Days to incident atrial fibrillation |
|  | Incident heart failure | Partial matching coding scheme and value levels. Action taken: We used cohort adjudicated data based on cohort determinations from hospitalization records, ICD codes, or self-report. Data not available in WHICAP. | Combined and recoded: Incident heart failure (yes/no) |
|  | Follow-up time to incident heart failure (definite or probable) | Complete coding matching. Action taken: In NOMAS, the follow-up interview date when CHF was reported was used to calculate days since cohort entry. Data not available in WHICAP. | Combined and recoded: Days to incident heart failure |
|  | Incident stroke of any type (definite/probable) | Partial matching in coding scheme and value levels. Action taken: We used the original cohort adjudicated variable and recoded to include only definite and probable incident cases, excluding those with events before study entry. Not available in WHICAP. | Combined and recoded: Incident stroke (yes/no) |
|  | Follow-up time to incident stroke (definite or probable) | Action taken: Follow-up time from cohort entry was standardized to days across cohorts; units were converted when needed. This variable was not available in WHICAP. | Combined and recoded: Days to incident stroke |
|  | Type of incident stroke (ischemic vs hemorrhagic) | Partial matching in coding scheme and value levels. Action taken: We used definite/probable criteria only, and combined and recoded stroke types to create ischemic, hemorrhagic or other/unknown incident stroke type variable. Not available in WHICAP. | Combined and recoded: Incident stroke type<br>Ischemic, Hemorrhagic, Other/unknown stroke |
|  | Type of incident hemorrhagic stroke (subtype) | Partial matching in coding scheme and value levels. Action taken: Combined and recoded variables in each cohort to create one 4 category incident hemorrhagic stroke subtype variable. Not available in WHICAP. | Combined and recoded: Incident stroke hemorrhagic subtype<br>Not hemorrhagic, SAH, IPH, Other/unknown hemorrhagic |
|  | Type of incident ischemic stroke (subtype) | Partial matching in coding scheme and value levels. Action taken: Variables were combined and recoded in each cohort to create a 5-category incident ischemic stroke subtype variable. Data not available in WHICAP. | Combined and recoded: Incident stroke ischemic subtype:<br>Not ischemic, Large vessel, Cardioembolic, Small vessel, Other/unknown ischemic |
| **Vital status and cause of death** |  |  |  |
|  | Death | Complete matching coding. Action taken: recoded response categories. | Recoded variable: Death (yes/no) |
|  | Follow up time to death | Complete matching variable. | Days to death |
|  | Atherosclerotic cardiovascular disease (ASCVD) death | Partial matching of coding scheme and value levels. Action taken: In ARIC, we used death certificate ICD-9 codes 390–459 and ICD-10 codes I00–I99 to recode the variable. In all other cohorts, we used adjudicated indicators of cardiovascular cause of death. Participants who were alive were coded as zero (no). | Combined and recoded: Cause of death, CVD (yes/no) |
|  | MI or CHD death (fatal MI) | Partial matching of coding scheme and value levels. Action taken: Fatal CHD/MI was identified using available variables indicating cause of death, including fatal MI, cardiovascular death codes, or atherosclerotic CHD. For all cohorts, participants who were alive were coded as no (zero). | Combined and recoded the variable: Cause of death, MI/CHD (yes/no) |
|  | Fatal stroke | Partial matching of coding scheme and value levels. Action taken: Fatal stroke was identified using ICD-9/10 codes from death certificates in ARIC (430-438, I60-69). In CHS, we combined fatal stroke and cardiovascular cause of death variables. In REGARDS, NOMAS, and MESA, we recoded variables indicating stroke death. In WHICAP, cerebrovascular cause of death was used. Participants who were alive were coded as zero (no). | Combined and recoded: Cause of death, stroke (yes/no) |

## Notes

### Competing Interest Statement

The authors have declared no competing interest.

### Funding Statement

Epilepsy-Cog: This work was supported by the National Institutes of Health (grant R01AG074355). Drs. Thacker, Choi, and Gutierrez were supported by this grant. ARIC: The Atherosclerosis Risk in Communities study has been funded in whole or in part with Federal funds from the National Heart, Lung, and Blood Institute, National Institutes of Health, Department of Health and Human Services, under Contract nos. (75N92022D00001, 75N923022D00002, 75N92022D00003, 75N92022D00004, 75N92022D00005). The authors thank the staff and participants of the ARIC study for their important contributions. CHS: This research was supported by contracts HHSN268201200036C, HHSN268200800007C, HHSN268201800001C, N01HC55222, N01HC85079, N01HC85080, N01HC85081, N01HC85082, N01HC85083, N01HC85086, 75N92021D00006, and grants U01HL080295, U01HL130114, and R01HL172803 from the National Heart, Lung, and Blood Institute (NHLBI), with additional contribution from the National Institute of Neurological Disorders and Stroke (NINDS). Additional support was provided by R01AG023629 from the National Institute on Aging (NIA). A full list of principal CHS investigators and institutions can be found at CHS-NHLBI.org. MESA: MESA is supported by contracts 75N92025D00022, 75N92020D00001, HHSN268201500003I, N01-HC-95159, 75N92025D00026, 75N92020D00005, N01-HC-95160, 75N92020D00002, N01-HC-95161, 75N92025D00024, 75N92020D00003, N01-HC-95162, 75N92025D00027, 75N92020D00006, N01-HC-95163, 75N92025D00025, 75N92020D00004, N01-HC-95164, 75N92025D00028, 75N92020D00007, N01-HC-95165, N01-HC-95166, N01-HC-95167, N01-HC-95168 and N01-HC-95169 from the National Heart, Lung, and Blood Institute, and by grants UL1-TR-000040, UL1-TR-001079, and UL1-TR-001420 from the National Center for Advancing Translational Sciences (NCATS). NOMAS: This project has been funded at least in part with federal funds from the National Institutes of Health, National Institute of Neurological Disorders and Stroke by R01-NS29993. REGARDS: This research project is supported by cooperative agreement U01 NS041588 co-funded by the National Institute of Neurological Disorders and Stroke (NINDS) and the National Institute on Aging (NIA), National Institutes of Health, Department of Health and Human Service. The content is solely the responsibility of the authors and does not necessarily represent the official views of the NINDS or the NIA. Representatives of the NINDS were involved in the review of the manuscript but were not directly involved in the collection, management, analysis or interpretation of the data. The authors thank the other investigators, the staff, and the participants of the REGARDS study for their valuable contributions. A full list of participating REGARDS investigators and institutions can be found at: https://www.uab.edu/soph/regardsstudy/. WHICAP: Data collection and sharing for this project was supported by the Washington Heights-Inwood Columbia Aging Project (WHICAP,R01AG072474, RF1AG066107, PO1AG07232, R01AG037212, RF1AG054023) funded by the National Institute on Aging (NIA). This manuscript has been reviewed by WHICAP investigators for scientific content and consistency of data interpretation with previous WHICAP Study publications. We acknowledge the WHICAP study participants and the WHICAP research and support staff for their contributions to this study

### Author Declarations

The Columbia University Institutional Review Board (IRB), which serves as the single IRB for Epilepsy-Cog, approved the use of de-identified data from the six cohorts for harmonization and pooling. IRBs of participating institutions involved in the six cohort studies approved the individual cohort studies.

